# Coverage and effectiveness of conditional cash transfer for people with drug resistant tuberculosis in Zimbabwe: a mixed methods study

**DOI:** 10.1101/2022.08.19.22278863

**Authors:** Collins Timire, Charles Sandy, Rashida A Ferrand, Regina Mubau, Peter Shiri, Obert Mbiriyawanda, Fredrick Mbiba, Rein MGJ Houben, Debora Pedrazzoli, Virginia Bond, Nicola Foster, Katharina Kranzer

## Abstract

**Background:** The End TB strategy recommends social protection to mitigate socio-economic impacts of tuberculosis. Zimbabwe started implementing a conditional cash transfer (CCT) programme for people on drug resistant tuberculosis (DR-TB) treatment in 2013. We aimed to determine the proportion of people receiving CCT and effectiveness of CCT in improving treatment outcomes, explore their experiences with registering for CCT and understand the impact of CCT from the perspective of beneficiaries.

**Methods:** Data from 2014-2021 were extracted from TB registers and CCT payment records within the National TB Programme. Sixteen in-depth interviews were conducted with people who were completing treatment or had completed treatment within two months. Poisson regression, adjusted for province, year of treatment, age and sex was used to investigate associations between receiving CCT and successful treatment outcomes among people who were in DR-TB care for ≥3 months after treatment initiation. Qualitative data were analyzed using thematic analysis.

**Results:** A total of 481 people were included in the quantitative study. Of these, 53% (254/481) received CCT at some point during treatment. People who exited DR-TB care within three months were 73% less likely to receive CCT than those who did not (prevalence ratio (PR)=0.27 [95%CI: 0.18-0.41]). Among those who were alive and in care three months after treatment initiation, CCT recipients were 32% more likely to have successful outcomes than those who did not (adjusted PR=1.32, [95%CI: 1.00-1.75]). Qualitative results revealed lack of knowledge about availability of CCT among people with DR-TB and missed opportunities by healthcare providers to provide information about availability of CCT. Delays and inconsistencies in disbursements of CCT were frequent themes.

**Conclusion:** CCT were associated with successful treatment outcomes. Improvements in coverage, timeliness and predictability of disbursements are recommended.

## Introduction

Only 59% of people receiving treatment for rifampicin resistant/multi-drug resistant tuberculosis (RR/MDR-TB) were reported to have successful treatment outcomes globally.(1) Zimbabwe mirrors the global picture as treatment success among people on DR-TB treatment has plateaued around 54%.(2,3) Successful treatment outcomes are defined as treatment completion and cure while unsuccessful outcomes comprise deaths, loss to follow-up, failure and relapses.(4)

Efforts to improve DR-TB treatment outcomes include i) earlier diagnosis ii) shorter and less toxic DR-TB regimens and iii) individualised regimens taking into account drug susceptibility results, as highlighted in the first and third pillars of the End TB strategy.(5) While shorter DR-TB regimens have improved treatment outcomes in some settings,(6) additional interventions aimed at mitigating the economic burden experienced by people affected by DR-TB may be needed.(5,7)

The pooled average of catastrophic costs, i.e. TB-related costs exceeding 20% of a household’s annual income prior to TB was estimated globally at 43% (95% CI: 34–51) among TB affected households. (8) This is much higher than the End TB strategy milestone of zero catastrophic cost by 2020. Major drivers of catastrophic costs are outside the health system and include food, transport, and income loss. Across settings, DR-TB affected households incur higher costs than households affected by drug-susceptible TB,(9–14) which may push households into financial catastrophes, and may act as barriers to accessing TB diagnostic and treatment services.(15) The End TB strategy acknowledges the need for greater action on the social determinants of TB through implementing universal health coverage and social protection.(5)

Social protection is an efficient, redistributive way to mitigate the socio-economic impact of chronic diseases.(16) It builds resilience of households and may improve attendance at health facilities and hence health seeking behaviour, which in turn may promote good health outcomes.(17,18) In the context of TB social protection may be TB inclusive, TB sensitive and TB specific.(19) Eligibility of TB inclusive schemes includes a diagnosis of TB or living with a household member with TB. TB sensitive interventions target people at high risk of TB through provision of basic income to reduce current and future poverty which in turn may protect people from TB. However, sustainability of TB sensitive interventions is challenging in resource limited settings.(19)TB specific interventions target people or households affected by TB and aim to improve specific TB indicators such as adherence and treatment outcomes.(19,20) TB specific social protection interventions may be conditional on beneficiaries adopting specific health behaviour such as regular clinic attendance or unconditional which come without obligations to beneficiaries.(21,22) The effectiveness of social protection depends on coverage, size of payouts and timeliness of disbursements.(17,23,24) Early disbursements are crucial since financial resources for most patients are exhausted by the time they are diagnosed with TB.(7,15)

Zimbabwe, a high HIV-TB and DR-TB burden country introduced a DR-TB specific conditional cash transfer (CCT) programme in 2013. The cash transfers are conditional on people attending follow-up appointments at health facilities. In practice the conditionality is not enforced. Eligibility for CCT is enrolment into DR-TB treatment regardless of socio-economic status of the person or household. Under the scheme, people with DR-TB receive USD 25 every month for the duration of treatment. Until December 2019, the cash was disbursed through a mobile money service called EcoCash. Thereafter, the cash was disbursed via bank platforms.

This study aimed to evaluate the programme with regards to coverage and effectiveness of CCT in improving treatment outcomes, using a mixed methods approach.

## Methods

### Study setting

The study was conducted in four provinces of Zimbabwe: Masvingo, Matebeleland South, Harare and Bulawayo. These provinces were purposively selected because of high DR-TB notification rates. In 2020, the estimated TB incidence was 193 per 100,000 population in Zimbabwe; prevalence of DR-TB was 3.1% among new cases and 14.0% among retreatment cases.(2,25) Prior to 2018, people with DR-TB received standardized 18-24 months regimens including an injectable drug, usually kanamycin.(26) In 2018, a 9-12 months all-oral regimen was implemented for those without resistance to quinolones.

### The conditional cash transfer programme for people with DR-TB in Zimbabwe

In 2013 the National TB Programme (NTP) introduced a financial support system of USD 25 per month for people with DR-TB. The programme aimed to disburse the first cash transfer within three months of starting treatment.

All people enrolled for DR-TB treatment are eligible for cash. The cash was initially disbursed in quarterly lump-sums of USD 75 using EcoCash a mobile money service. In June 2017 Zimbabwe introduced a local currency (ZWL) which initially traded at 1:1 against the USD. EcoCash could no longer offer USD, but used an in-built conversion factor based on the prevailing interbank exchange rate to convert USD into local currency. By April 2020 and February 2022, USD 1 was equivalent to ZWL 25 and ZWL 124, respectively against the official interbank exchange rate.(27) The unofficial (black market) rate is usually higher than the interbank rate. Cash was chosen instead of food and food vouchers because it is logistically easier to distribute and can be accessed even in remote areas. Cash also gives people affected by DR-TB options with regards to the mix and type of products and services (e.g. food and transport to health facilities) they can purchase.(16)

From 2014 to 2019, the money was disbursed through a mobile money service (phone-based payment) called EcoCash, enabling customers to receive CCT and transact directly on mobiles phones. A national identity document was required to open an EcoCash account, which some did not have. People affected by DR-TB who could not meet these requirements were asked to use EcoCash accounts of their next of kin or third parties. Of note, EcoCash connectivity is suboptimal in some areas including Matebeleland South province. The method of payment was changed in January 2020 after EcoCash failed to honour their obligation to pay USD as alluded above. This prompted the NTP to migrate all the patients to bank payment platforms in four commercial banks that covered all geographical areas.

The processing of cash transfers involves multiple tiers of the health care system and is entirely paper-based. At the end of each month, health facilities compile a list of names of people receiving DR-TB treatment. At district level, all lists from health facilities are consolidated, signed by the District Medical Officer and are submitted to the Provincial Medical Directorate for consolidation into a provincial return before they are forwarded to the national (central) office. The NTP removes duplicate names and people who completed treatment, were lost to follow-up or had died. A payment authorization is submitted to the finance department within the Ministry of Health and Child Care (MoHCC) for review and to the NTP Manager for approval. Final approval is sought from the Director of Finance MoHCC before cash is transferred into EcoCash or bank accounts.

### Study population and data collected for the quantitative study

All people with DR-TB who started treatment between 01 January 2014 and 31 January 2021 in 35 selected health facilities in Masvingo, Matebeleland South, Harare and Bulawayo province were eligible for inclusion in the study. Data were extracted from TB treatment registers and CCT disbursement logs using a structured questionnaire. The following variables were collected: age, sex, HIV status; type of TB, province; date of enrolment on treatment; treatment outcomes and date of outcome. Centrally held CCT log sheets were used to determine whether or not a person received cash transfers, when the first cash transfer was disbursed and total amount disbursed.

### Study population and data collection for qualitative study

Sixteen adults (≥18 years) living in Harare (urban) and Matebeleland South (rural) provinces who had either completed DR-TB treatment within two months or were left with less than two months to complete treatment were purposively selected (taking account of age, sex and place of residence) to take part in in-depth interviews. All interviews were audio-recorded and were conducted in local languages (Shona and Ndebele) in private locations suggested by participants. Interview themes included: illness trajectory, health seeking behaviour, knowledge about availability of CCT, experiences with getting registered for CCT, timeliness of CCT, how the CCT helped and suggestions to policy makers about CCT. Interviews were transcribed and translated into English.

### Data Analysis

Data were entered in EpiData v3.1 (EpiData Association, Odense, Denmark) and were exported to STATA v17 (StataCorp, College Station, TX) for analysis. The amount of CCT disbursed was used to categorise people with DR-TB into having received at least one CCT or not having received a CCT (exposure). Continuous data e.g. age were assessed for normality using the Shapiro-Wilk test and were summarized using means and standard deviations. Categorical data were summarized using frequencies and proportions. The proportion of people receiving at least one CCT was calculated by province, year of treatment, age, sex HIV status, treatment regimen and retreatment status. The factors associated with receiving CCT were investigated using binominal regression.

Treatment outcomes were categorized into successful (completed treatment and cured) and unsuccessful (died, failed treatment and lost to follow-up). The effect of CCT (exposure) on treatment outcomes (outcome) was investigated using binomial regression in a cohort of patients who were alive and in DR-TB care at three months post-treatment initiation as this was the minimum time it took to disburse the first CCT. Prior to multivariable analysis, a directed acyclic graph (DAG) (**Supplementary Figure 1)** was drawn using DAGitty (http://www.dagitty.net/) to identify confounding variables. Poisson regression with robust variance estimators was then performed to investigate the effect of CCT on treatment outcomes adjusting for province, year of treatment, age and sex (confounders identified by the DAG).

Qualitative data were coded both manually and using *NVivo* version 12 (QSR International). Codes were data and concept driven. Codes were grouped into themes during an iterative process. Data were analyzed using thematic analysis.(28) Names used in quotes have been pseudonymized.

#### Ethics

Ethical approval was obtained from the London School of Hygiene and Tropical Medicine Research Ethics Committee (22579), the Biomedical Research and Training Institute (AP160/2020) and the Medical Research Council Zimbabwe (MCRZ/A/2645). Permission to access DR-TB records was obtained from the Secretary for Health, Ministry of Health and Child Care Zimbabwe. Written informed consent to take part in and to audio-record in-depth interviews was obtained from all participants.

## Results

Quantitative data for 481 people were extracted. Their mean age was 37.6 years (standard deviation 11.9). The majority (n=292, 61%) were men and 369 (77%) were co-infected with HIV (Table 1). Bulawayo and Matebeleland South provinces contributed the majority of participants (n=305, 64%). Most (n=380, 79%) had been on an 18-24 month regimen.

**Table 1:**
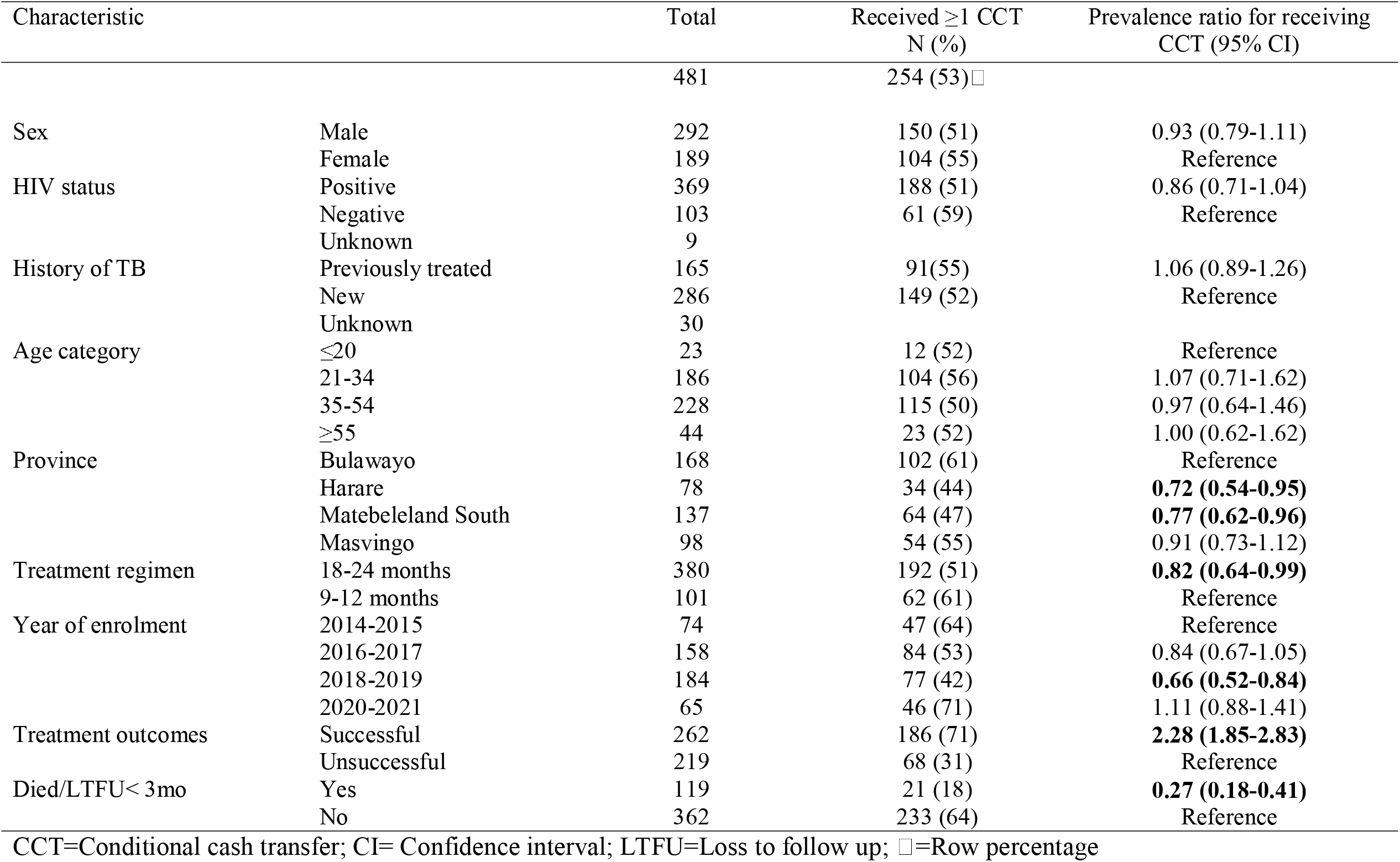
Demographic, clinical characteristics, treatment outcomes and receipt of CCT of people enrolled in the study, 2014-2021, Zimbabwe

| Characteristic | | Total | Received $\geq 1$ CCT<br>N (%) | Prevalence ratio for receiving<br>CCT (95% CI) |
| --- | --- | --- | --- | --- |
|  |  | 481 | 254 (53) □ |  |
| Sex | Male | 292 | 150 (51) | 0.93 (0.79-1.11) |
|  | Female | 189 | 104 (55) | Reference |
| HIV status | Positive | 369 | 188 (51) | 0.86 (0.71-1.04) |
|  | Negative | 103 | 61 (59) | Reference |
|  | Unknown | 9 |  |  |
| History of TB | Previously treated | 165 | 91 (55) | 1.06 (0.89-1.26) |
|  | New | 286 | 149 (52) | Reference |
|  | Unknown | 30 |  |  |
| Age category | $\leq 20$ | 23 | 12 (52) | Reference |
|  | 21-34 | 186 | 104 (56) | 1.07 (0.71-1.62) |
|  | 35-54 | 228 | 115 (50) | 0.97 (0.64-1.46) |
| | $\geq 55$ | 44 | 23 (52) | 1.00 (0.62-1.62) |
| Province | Bulawayo | 168 | 102 (61) | Reference |
|  | Harare | 78 | 34 (44) | **0.72 (0.54-0.95)** |
|  | Matebeleland South | 137 | 64 (47) | **0.77 (0.62-0.96)** |
|  | Masvingo | 98 | 54 (55) | 0.91 (0.73-1.12) |
| Treatment regimen | 18-24 months | 380 | 192 (51) | **0.82 (0.64-0.99)** |
|  | 9-12 months | 101 | 62 (61) | Reference |
| Year of enrolment | 2014-2015 | 74 | 47 (64) | Reference |
|  | 2016-2017 | 158 | 84 (53) | 0.84 (0.67-1.05) |
|  | 2018-2019 | 184 | 77 (42) | **0.66 (0.52-0.84)** |
|  | 2020-2021 | 65 | 46 (71) | 1.11 (0.88-1.41) |
| Treatment outcomes | Successful | 262 | 186 (71) | **2.28 (1.85-2.83)** |
|  | Unsuccessful | 219 | 68 (31) | Reference |
| Died/LTFU < 3mo | Yes | 119 | 21 (18) | **0.27 (0.18-0.41)** |
|  | No | 362 | 233 (64) | Reference |

CCT=Conditional cash transfer; CI= Confidence interval; LTFU=Loss to follow up; □=Row percentage

Overall, 254 (53%) of participants received at least one cash disbursement (Table 1). The proportion receiving CCT varied across time and province with the highest proportions recorded in 2020-2021 (71%) and in Bulawayo province (61%) (Table 1 and Figure 1). Successful treatment outcomes were recorded for 262 (54%). A quarter 119 (25%) died or were lost to follow up (LTFU) within three months of starting treatment (Figure 2, Table 2, Supplement Figure 2). Of these 119 people, only 21 (18%) received CCT.

**Figure 1:**
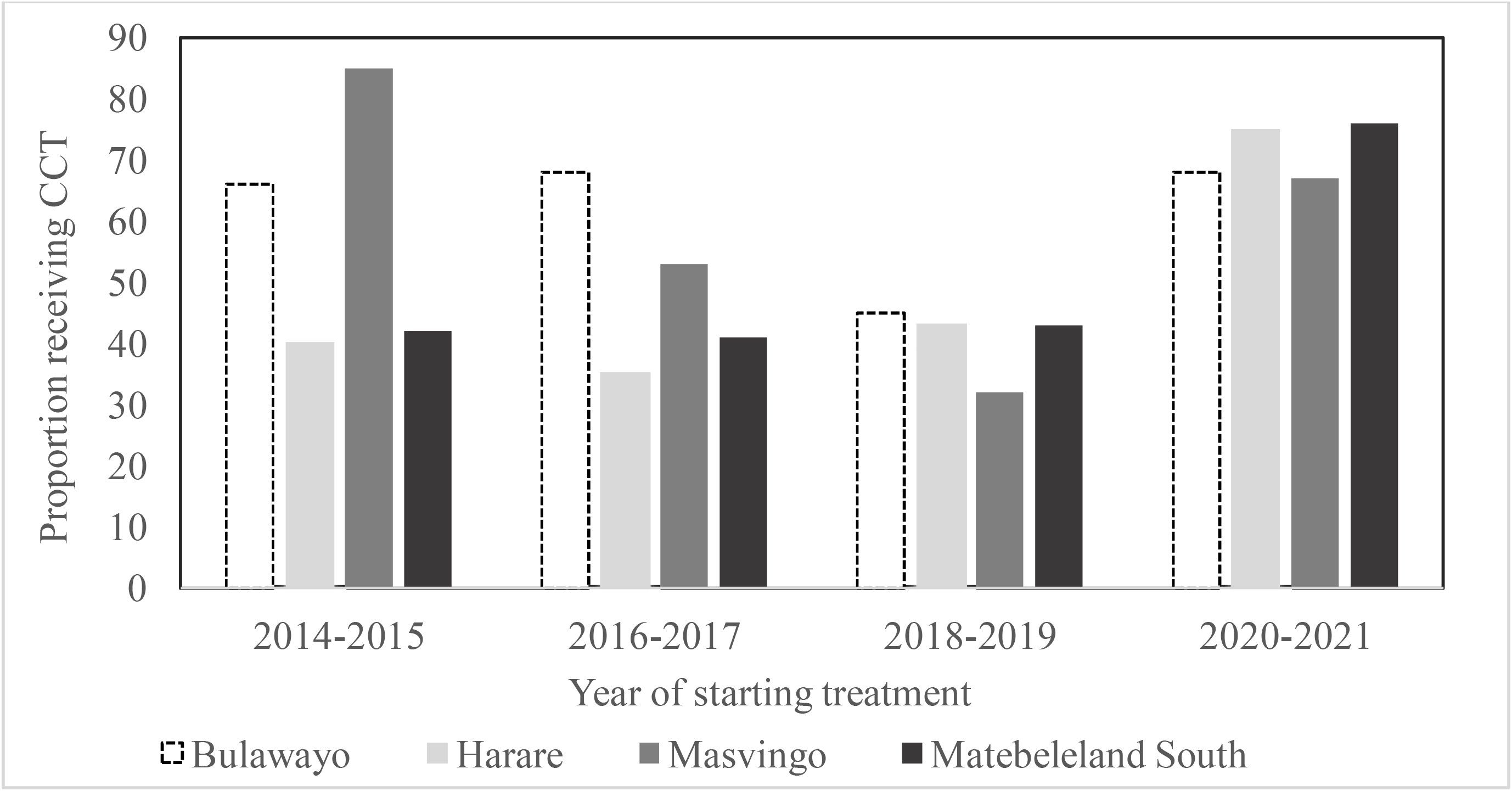
Proportion of people with DR-TB who received CCT by province and year of enrolment, 2014-2021, Zimbabwe

**Figure 2:**
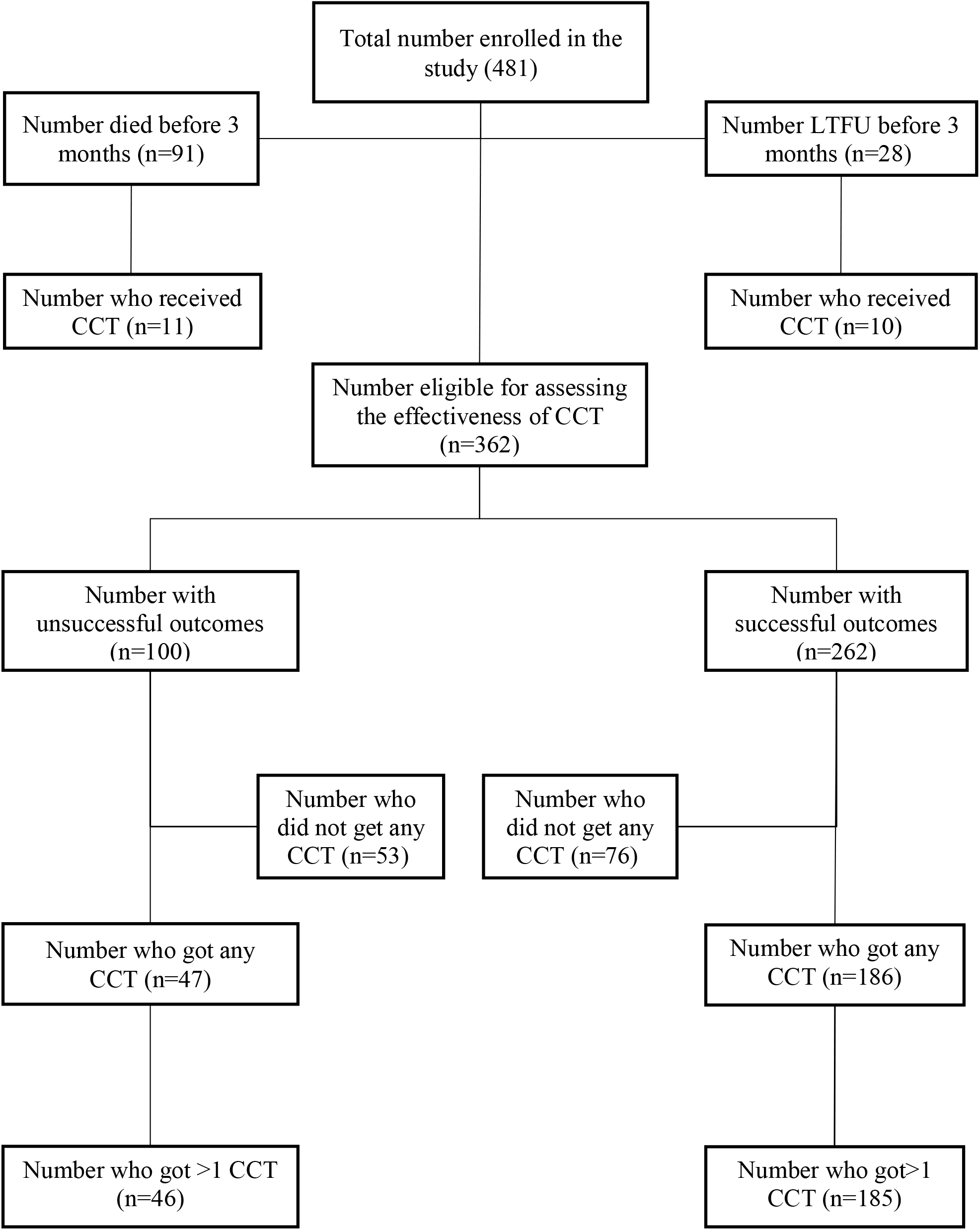
Flowchart for people with DR-TB whose records were included in the study, Zimbabwe, 2014-2021

**Table 2:** Factors associated with successful outcomes among people with DR-TB who were enrolled in the study, Zimbabwe, 2014-2021

| Characteristic |  | Total | Successful outcomes<br>Number (%)† | PR (95% CI) | aPR (95% CI) |
| --- | --- | --- | --- | --- | --- |
| Total |  | 362 | 262 (72) |  |  |
| Sex | Male | 221 | 156 (71) | 0.94 (0.83-1.07) | 0.95 (0.74-1.23) |
|  | Female | 141 | 106 (75) | Reference | Reference |
| Age category (years) | ≤20 | 18 | 13 (72) | Reference | Reference |
|  | 21-34 | 147 | 110(75) | 1.04 (0.77-1.40) | 1.05 (0.59-1.88) |
|  | 35-54 | 172 | 123 (72) | 0.99 (0.73-1.34) | 1.02 (0.57-1.84) |
|  | ≥55 | 25 | 16 (64) | 0.89 (0.59-1.34) | 0.90 (0.43-1.89) |
| Province | Bulawayo | 126 | 100 (79) | 1.07 (0.91-1.25) | 1.05 (0.75-1.47) |
|  | Harare | 65 | 44 (68) | 0.91 (0.73-1.13) | 0.96 (0.64-1.44) |
|  | Matebeleland South | 97 | 63 (65) | 0.87 (0.71-1.07) | 0.88 (0.61-1.28) |
|  | Masvingo | 74 | 55 (74) | Reference | Reference |
| Year of enrolment | 2014-2015 | 58 | 41 (71) | Reference | Reference |
|  | 2016-2017 | 113 | 80 (71) | 1.00 (0.82-1.23) | 1.03 (0.72-1.55) |
|  | 2018-2019 | 137 | 95 (69) | 0.98 (0.80-1.20) | 1.08 (0.74-1.60) |
|  | 2020-2021 | 54 | 46 (85) | 1.21 (0.99-1.47) | 1.21 (0.79-1.86) |
| Registered for CCT | Yes | 233 | 186 (80) | **1.35 (1.16-1.59)** | **1.32 (1.00-1.75)** |
|  | No | 129 | 76 (59) | Reference | Reference |

†=Row percentage; PR=prevalence ratio; aPR=adjusted prevalence ratio; CCT=Conditional cash transfer; CI=confidence interval

### Effectiveness of CCT on treatment outcomes

The NTP aims to disburse the first CCT within three months of starting treatment. Hence the analysis investigating the effectiveness of CCT in treatment outcomes was assessed on a cohort of 362 people who were alive and in DR-TB care at 3 months post treatment initiation − the earliest time they could have received CCT. Of the 362 people, 262 (72%) had successful outcomes (Figure 2 and Table 2). In univariate analysis, there was a strong evidence for association between receipt of CCT and successful outcomes. Those who received CCT were 35% more likely to have successful treatment outcomes than those who did not, (prevalence ratio (PR) =1.35 [95%CI: 1.16-1.59]) (Figure 3, Table 2). Adjustment for age, sex, province and year of treatment did not change the prevalence ratio substantially, (aPR=1.32, [95% CI 1.00-1.75])

**Figure 3:**
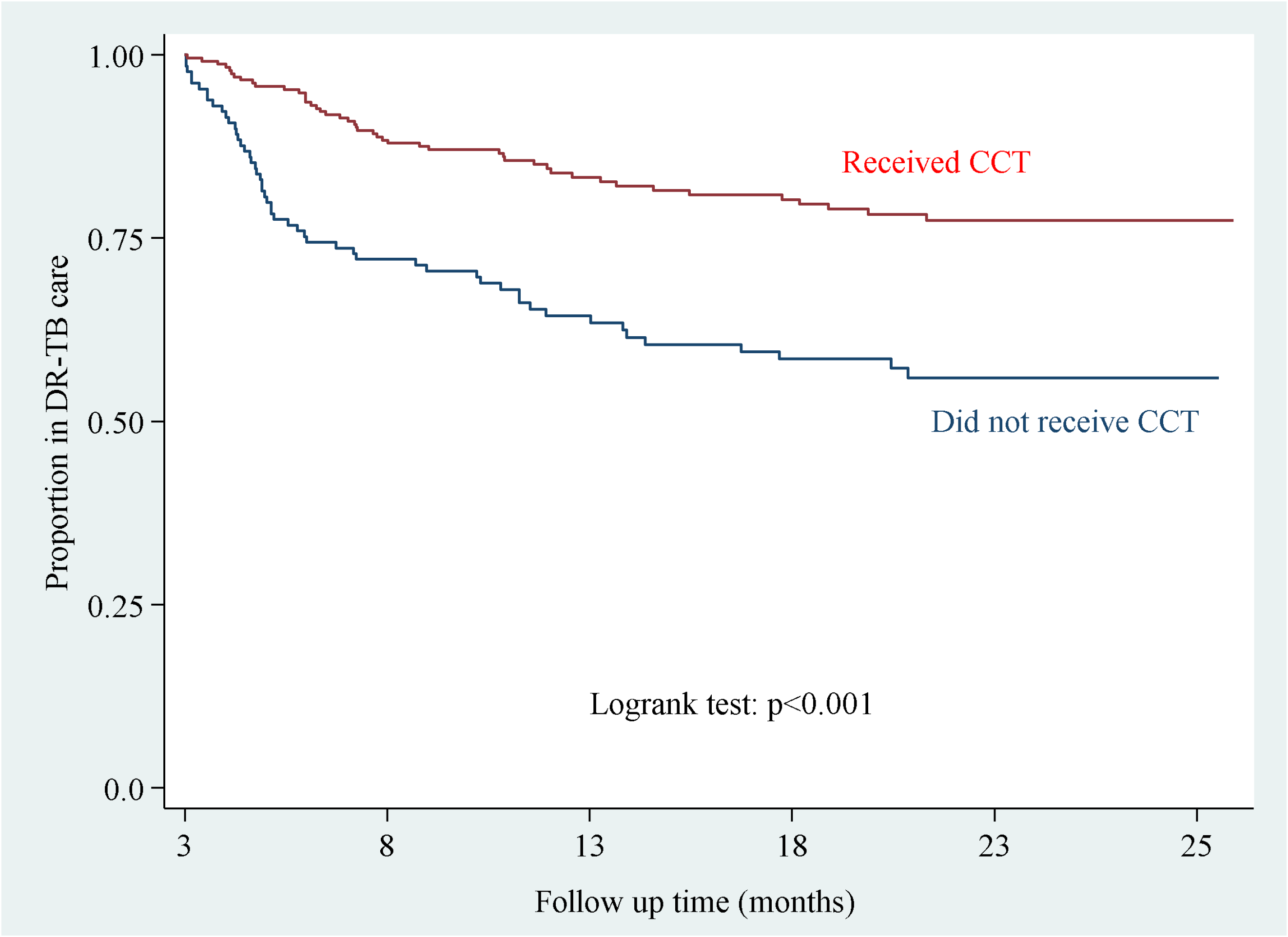
Time to unsuccessful outcomes among people with DR-TB, by registration status for CCT, 2014-2021, Zimbabwe

### Lack of knowledge about the availability of CCT

People with DR-TB viewed DR-TB as a unique disease which required that they be supported in order for them to complete treatment. However, none of the participants knew about the availability of CCT prior to getting registered (Table 3). For some, lack of knowledge about availability of CCT persisted until they completed treatment. Registration for CCT was only achieved if it was initiated by healthcare workers.

**Table 3:** Themes and quotes.

| Themes | Quotes of supporting themes |
| --- | --- |
| Lack of knowledge about availability of CCT | *Uhmm what you are saying is new to me. For two and a half years I never heard of such a thing that there is money [chuckles] that is supposed to be given to people. Peter_M‡* |
|  | *Uum. I didn't know about the incentives. When we went to the clinic there was a certain man who came requesting for our names and other details so that we would be given some money. Ruth_F* |
| Lack of knowledge about registration status | *No. I haven't received it. They just asked me to send my ID number. I sent it but nothing happened after that. I didn't receive money. Spiwe_F* |
| Easy registration process | *It [registering for CCT] wasn't difficult for me. Agnella_F* |
| CCT may not provide enough cushion against income loss | *Considering everything that we experienced, the money is too little even without factoring in the money we spent at traditional healers. Traditional healers charge exorbitant fees as compared to hospitals. If you go to hospitals, you only pay for transport since treatment is free, but at traditional healers, you can't leave the place without paying something. That's how they operate. Caleb_M* |
|  | *It helped me on food but I couldn't afford to pay rentals with it. Agnella_F* |
| Delays and unpredictability of CCT | *It surely came but as for now it has been long since I last received the money because it came once. Moses_M* |
|  | *Aaah the money took some time before it came. It comes here and there. It doesn't come monthly. Sometimes it comes and then stops. Like the last batch was in December. Loveness_F* |
|  | *They should disburse the money on time. Even if it's little but it should be disbursed on time. Loveness_F* |
| | *Even if it is \$10 or \$1 after 5 days. Let the \$1 come after 5 days not to be told that the money has finally come after 9 months! Caleb_M* |
| Poor connectivity as a barrier to registration | *So I just said to myself that, 'Aaahh, maybe since I stay where there are network challenges, maybe I delayed in sending the [ID] number to the programme. But I am wondering what happened since I did not get any feedback. I just concluded that maybe I delayed. Spiwe_F* |
| Post-TB sequeale | *I still don't have enough strength since I spent the whole year and some months on the sick bed. So am looking forward to gradually re-gain strength. Loveness_F* |
|  | *I managed to finish my medication even though I have some side effects, I experienced lots of side effects on my legs...they are numb such that I can't walk. For me to walk long distances.... Agnella_F.* |

‡= pseudonym and sex of participant

> *As for me, I didn’t know that there is financial assistance. I thought it [DR-TB] is like any other disease in which a man is for himself. But I have discovered that one cannot manage on his own. Moses_M*

Often, nurses were reported to be too focused on their core business of treating patients and dispensing medicines to consider the financial wellbeing of patients. This may have reduced chances for people to get registered for the CCT scheme.

> *The mistake that I have discovered is that, when a TB patient goes to hospital, their [nurses] only concern is to treat the patient. Ruth_F*

### Implementation challenges of the CCT programme

Delays in registration were worse for people who were diagnosed in private facilities and those initially started on first-line TB treatment. Also, people who were transferred from one facility to another experienced delays in registration. The following quote reflects registration assistance for one participant.

> *…then after some time when I was halfway into my treatment…That’s when the EHT [Environmental Health Technician] helped me with the paperwork. He also helped me with everything that was needed for me to have an EcoCash account. Godfrey_M*

Transfers mostly happened during the early stages of DR-TB treatment at a time when people rely on support by family and friends. Likelihood of CCT registration was also affected by disruptions due to COVID-19. Staff was redeployed to support COVID-19 efforts and to clinics which experienced staff shortages. This in turn impacted on the continuity of TB service provision.

> *We knew each other very well with the nurse who was there and was supposed to administer my injections the week that followed. But that nurse did not come to work because she has been transferred and there was a new nurse. I had to start explaining to her all over again. Peter_M*

The actual registration process was considered easy. However, once registration was completed there was often a long delay before the first cash transfer was received.

> *It didn’t take me long [to get registered] and he also advised that it might take 2 to 3 months for me to get the money but I was not supposed to lose hope since it was definitely going to come. Moses_M*

Inconsistency and delays of CCT disbursements were a recurrent theme. Delays in receiving CCT increased when EcoCash changed the disbursement conditions, prompting the NTP to switch to bank payment platforms. The switch, combined with the COVID-19 pandemic resulted in up to 12 months delays of CCT payouts in 2020.

> *There is a time we had problems with EcoCash. That’s when it was stopped again and we were asked to open accounts with ZB bank. Loveness_F*

> *I got registered in April and got it in February. So you see! From April going to X [hospital] everyday*…*April 2020 to February 2021…I got the money in February. It was USD200 but I got USD185. Caleb_M*

Delays in CCT disbursements coincided with periods when financial resources of most people were depleted. Further delays were experienced because people were not informed (through short message services) when money was credited to their accounts. Thus, additional costs were incurred because people had to travel to banks to check if their accounts had been credited. As a result of both delays and unpredictability of CCT, some were pushed into debts.

> *We had spent all the money that we had by the time I went back to the doctors at Wilkins who confirmed that it was TB. Moses_M*

> *It [money] helped us a lot but sometimes it would not come. So we ended up borrowing and by the time we got it, we were in debt. Agnella_F*

Delayed disbursements were backdated and were received as lump-sums which were helpful when finally received. Unlike the EcoCash platform, bank accounts store the value of CCT as USD. On the downside people with DR-TB incurred exorbitant bank charges. Also bank officials sometimes tried to pay out in local currency instead of USD.

> *The third disbursement was USD250…The third one, because they had opened banks accounts for us, we got it in hard currency but the bank gave us USD235*.*Tecla_F*

> *Then they [bank tellers] said, “So that’s the money you have. We are going to transfer it to your card [in local currency] then you can go and swipe for groceries”. When the bank manger overheard this, he said, “Give him USD!” I was given the money and I left. That’s how I got it. Caleb_M*

### Perceived impact of CCT from the perspective of beneficiaries

People had different perceptions on the usefulness of CCTs. Those who did not experience significant income loss or diagnostics delays found CCTs helpful. CCTs allowed to cover additional TB-related expenses and to purchase food to share with family and household members. When CCTs were delayed and received as lump-sums, debts could be repaid and financial obligations which were not directly related to survival such as schooling could be fulfilled.

> *Aaah and the money that I received from the Ministry [of Health] helped me a lot. Felistus_F*

> *It helped me a lot because that time I managed to lessen the burden on my parents who were taking care of me. I managed to buy some food stuffs, clothes for my children and school uniforms and books. Ruth_F*

By contrast, people who either incurred huge income loss and/or costs felt that CCTs were insufficient. Physical debility and lost income as a result of DR-TB, often exacerbated by late diagnosis, made the CCT disbursements inadequate to cover financial needs.

> *The injection itself causes dizziness. There is no way you can go back to work after getting the injection and a handful of tablets. The moment you start DR-TB treatment, your career is destroyed. Ruth-F*

> *Yes it helps. It cannot address every need but it helps. To tell the truth…the situation that I was in ….I needed to buy some sugar, cooking oil and everything since I am someone who had no income. So the money was little. Loveness_F*

Health challenges related to DR-TB are not only confined to the treatment period. Some people experienced health problems and physical disability even after completing DR-TB treatment. They continued to incur out of pocket costs for health and income loss since they were not able to work. However, CCT disbursements are stopped at the end of treatment.

> *Like now, I have problem with my legs, these toes shiver on their own. You see that? I don’t know what’s causing that. I also have numbness in my joints… I would like to go and see a doctor but I don’t have money. I need medical attention and food. Loveness_F*

### Suggestions provided by people on DR-TB treatment for improving CCT

People affected by DR-TB suggested an opt-out CCT registration system both at private and public institutions.

> *They must register the patient immediately wherever MDR is detected either at the hospital or private doctor. Moses_M*

Electronic patient management systems were suggested as a means to ease monitoring of CCT registration status of all people in DR-TB care even if they are transferred to other facilities.

> *Why is the information not in the system so that you access it automatically and you will be able to know that a-ah here we have this patient? Peter_M*

Timely and consistent CCT were highlighted as being important so that people with DR-TB are cushioned against economic hardships. This would help to plan expenditure and avert harmful coping strategies such as selling of assets and taking loans at exorbitant interest rates.

> *The problem is they fail to live up to their promise of releasing the funds at the stipulated dates as a result we suffer. Tecla_F*

> *We sold some chickens to get money to buy sugar, cooking oil or rice. So that’s how we survived until we reached a point where she would sell some spanners. She would look around the house to find out if there was something to sell so that we could get some money for bus fare. Moses_M*

## Discussion

This study found that receiving CCT increased the likelihood of successful treatment outcomes in those alive and remaining in DR-TB care at three months post treatment initiation. Only half of eligible individuals received CCT, with the majority experiencing over 10 months of CCT delays, especially during the COVID-19 period. A quarter of participants either died or were lost to follow within three months of starting treatment, the majority had not received any CCT. Death was the cause in 76% of those who exited the cohort during the first three months.

Most CCT schemes in the context of TB aim to improve treatment outcomes (microbiological cure and treatment completion).(23,29–31) The Zimbabwean National TB Strategic Plan 2021-2025 aims to improve DR-TB treatment success from 57% to 75% by 2025, mainly using innovative adherence support systems like CCTs. Evidence on the effect of social protection on TB treatment outcomes among people affected by DS- and DR-TB is still growing. One may argue that TB treatment outcomes measured for surveillance purposes are not the best metric to measure the effect of CCT. Measurements of wellbeing and protection against harmful coping strategies may be more meaningful. However, given that most CCT schemes aim to improve treatment outcomes, the results of this study showing that a positive effect of CCT among those who were in care and alive at three months are important. This is despite the qualitative results highlighting that USD 25 per month is insufficient to cover expenses and make up for lost income for most people affected by DR-TB. It is possible that the effect of CCT may have differed by socioeconomic status at the start of treatment. Data on household assets and income is not routinely captured in DR-TB registers and could not be considered in the analysis. However, the recent cost survey conducted in Zimbabwe, showed that 90% of DR-TB affected households experience catastrophic costs, highlighting the vulnerability and low socioeconomic status of most of these households.(11)

In line with this study, studies conducted in Nigeria, Brazil and Argentina reported positive effects of social protection on treatment outcomes.(29,30,32,33) By contrast, studies from South Africa and Timor-Leste showed either weak or no effect of social protection on adherence and/or treatment outcomes.(23,31) Possible explanations of these conflicting results could be heterogeneity in study designs, intervention packages and coverage of interventions. For example, a qualitative process evaluation of a trial investigating the effect of food vouchers on cure rates among people affected by TB in South Africa revealed that the limited effect may be explained by under-coverage and inconsistent payouts.(34)

Qualitative results of this study revealed frequent delays in receipt of CCT, a finding also reported in India and South Africa.(35,36) Delays in the Zimbabwean CCT scheme were worsened when the mode of disbursement was switched from EcoCash to bank accounts. Coverage of CCT was modest and comparable to findings from South Africa.(23) High attrition due to death and loss to follow-up within the first three months may explain some of the low coverage. Even among people who stayed in care beyond three months, only 64% received CCT. Transfers to other health facilities and high staff turnover were identified as hurdles for people to be registered for CCT. Additionally, healthcare workers were reported to be focused on administering medication rather than ensuring people affected by DR-TB were registered for the social protection they were eligible for. Critically, people with DR-TB were not aware about availability of CCT and were thus not empowered to demand social protection. Poor access, despite eligibility for social protection is a missed opportunity to improve the socio-economic situation of DR-TB affected households.(37,38) For better coverage, beneficiaries need to be linked to social protection more effectively.(39)

This study is strengthened by the fact that it was conducted under routine programme conditions. It therefore gives a real-world view of what is unfolding in the programme. Selection bias was minimised by including all the DR-TB records that were available in sampled facilities. Quantitative and qualitative data were triangulated to provide nuanced descriptions of participant experiences. However, this study has limitations. Firstly, some of the DR-TB registers in Harare (number could not be ascertained) could not be located. However, missing records were likely to be missing at random. Secondly, a key variable, the date of receipt of first CCT was not captured by the NTP. The date was estimated through use of proxy dates when CCT returns were processed by the finance department at the NTP. Because the exact date of disbursement was not available we did not perform an analysis investigating the effect of timely CCT on treatment outcomes. Thirdly, residual confounding cannot be ruled out since variables such as socio-economic data are not captured in TB registers. The qualitative data suggests that socio-economic status may be an effect modifier in the association between CCT and TB treatment outcomes. However, given the low socio-economic status of households affected by DR-TB, heterogeneity with regards to income and socioeconomic status is likely to be limited.(11) Also to account for delays in CCT disbursement we limited the analysis to those in care at 3 months of DR-TB treatment. By doing so we introduced survival bias which may have resulted in overestimating the effectiveness of CCTs. Lastly, we could not access the proportion of people whose returns were rejected on the basis of wrong account details. On average, around 13% of the returns are rejected (personal communication). It is not clear whether people with rejected returns access CCT at a later point in time. This means there may be a degree of misclassification in the exposure variable (receipt of CCT).

Despite these limitations, our study shows that CCT was effective even in a challenging operational environment, and highlights several areas for improvement/further strengthening. Firstly, rapid registration and expedited disbursement of the first installments are critical. This can be complemented with an account verification system at registration to help reduce number of rejected returns. Secondly, decentralising CCT disbursements to district level where patients are seen monthly will reduce administrative delays and facilitate earlier and more consistent disbursements. Also increased coverage may be achieved by revising DR-TB registers to capture data on CCT (registration status and receipt) and by embedding CCT in health promotion messaging provided by nurses to raise awareness about CCT among people with DR-TB. Monitoring of both coverage and timeliness of CCT could be enhanced through implementing electronic health records which are linked to CCT eligibility and disbursement records. Once a person is enrolled in DR-TB care and treatment duration is ascertained, the predictability and timeliness of CCT may be enhanced by ensuring that “disbursing CCT” becomes the default option. In light of a high prevalence of TB sequelae post DR-TB treatment and the associated income loss observed in this study and elsewhere, (40) extending CCT beyond treatment completion could be considered. Mitigation measures are needed to protect people and households against the socio-economic impacts of TB, including during periods of global shocks such as COVID-19. Programmes may increase the amount of CCT disbursed (vertical expansion) to mitigate extra shocks.(17) In 2022, the Zimbabwean NTP started complementing CCT with monthly grocery vouchers worth USD 20 for households affected by DR-TB.

## Conclusion

CCT were associated with successful treatment outcomes. Improvements in coverage, timeliness and predictability of disbursements are recommended.

## Data Availability

Data will be provided by the corresponding author upon request

## Acknowledgements

We would like to thank the participants who took part in this study and provided recommendations to improve the cash transfer for people with DR-TB in Zimbabwe.

## Author contributions

Conceptualised the study: CT, KK, DP, NF,VB, CS

Reviewed the draft manuscript: CT, KK, DP, VB, RMGJH

Data collection: CT, FM, OM, RM, PS

Data analysis: CT, KK, VB

Critical review of manuscript: CT, CS, KK, VB, RMGJH, RAF, NF, OM, PS, RM, FM, DP.

All authors reviewed and approved the final manuscript.

## Ethics statement

Ethical approval was obtained from the London School of Hygiene and Tropical Medicine Research Ethics Committee (22579), the Biomedical Research and Training Institute (AP160/2020) and the Medical Research Council Zimbabwe (MCRZ/A/2645). Permission to access DR-TB records was obtained from the Secretary for Health, Ministry of Health and Child Care Zimbabwe. All participants who took part in in-depth interviews gave written informed consent to take part in the study and to use an audio-recorder.

---

**Supplementary Figure 1:**
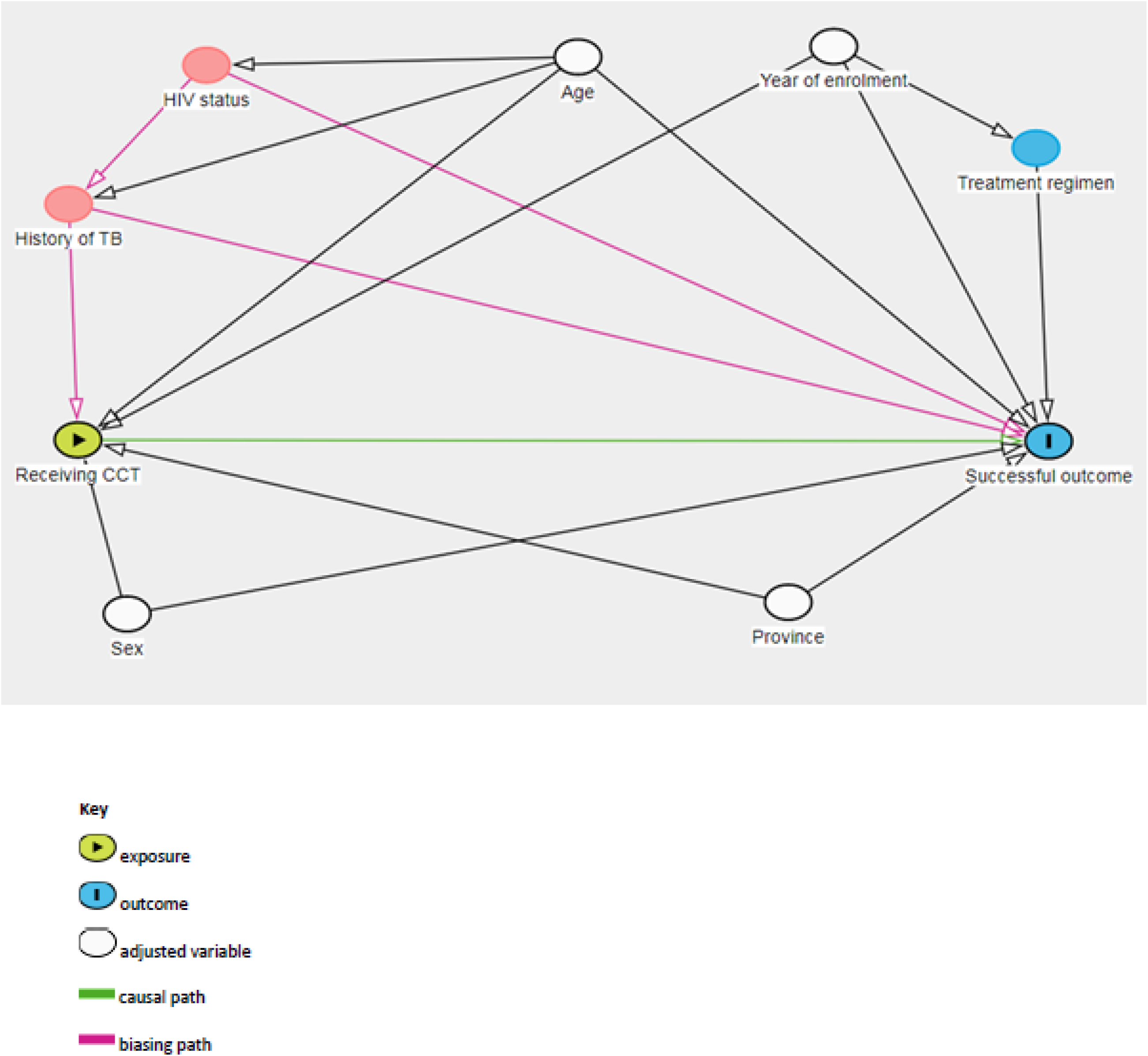
Causal diagram showing relationship between receipt of CCT and treatment outcomes with potential confounders.

**Supplementary Figure 2:**
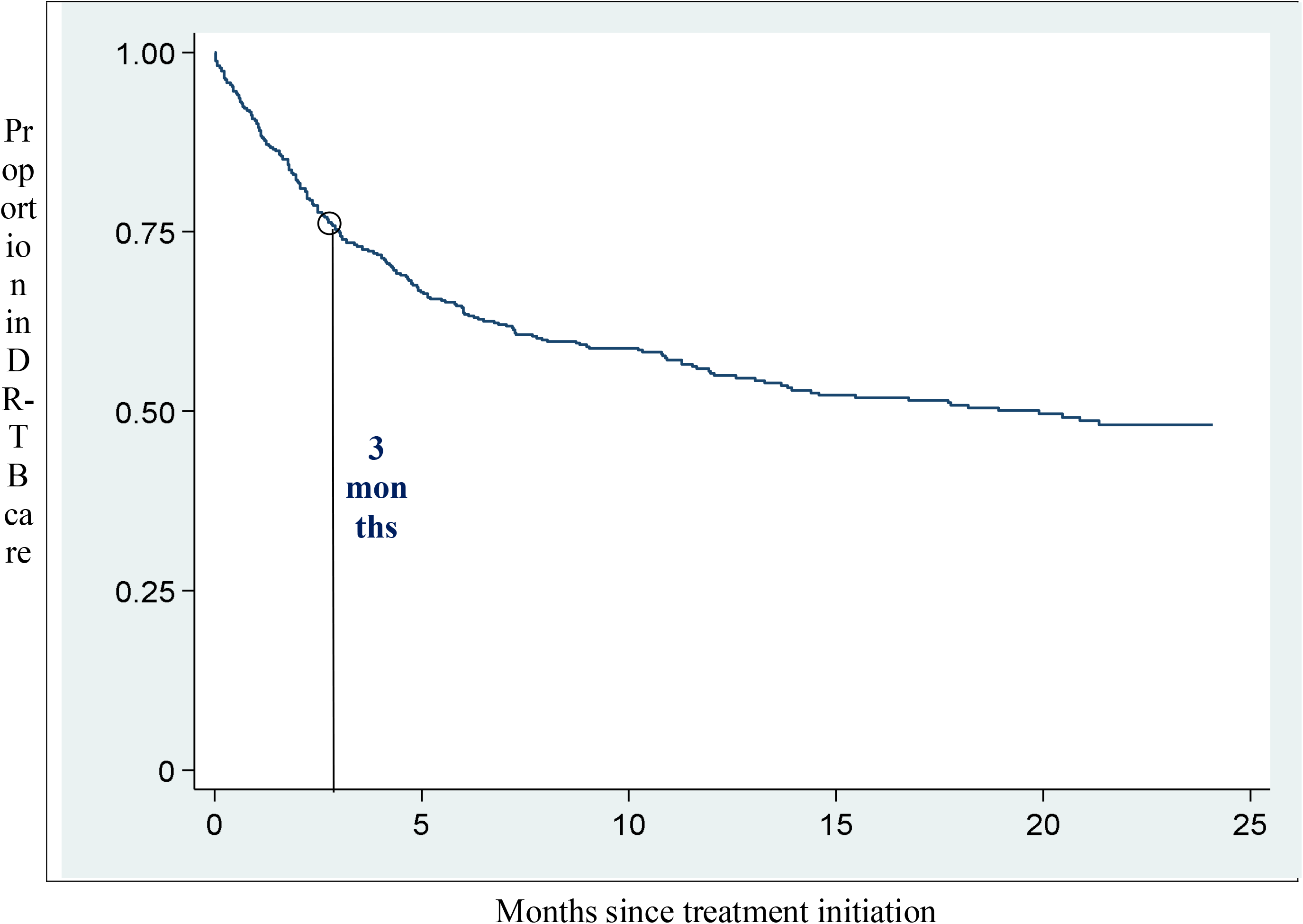
Time to unsuccessful outcomes among people with DR-TB

**Supplementary Table 1:** Demographic and clinical characteristics of DR-TB participants who were enrolled in the qualitative interviews, Zimbabwe, 2020-2021

| Study ID | HIV status | Duration of treatment | Marital status | History of TB | Formally employed | Received CCT |
| --- | --- | --- | --- | --- | --- | --- |
| Agnella_F‡ | Positive | 18-24mo | Single mother (2 children) | No | No | Yes |
| Ruth_F | Positive | 9-12mo | Single mother | Yes. | No | Yes |
| Felistus_F | Negative | 18-24mo | Single, never married | Yes | No | Yes |
| Moses_M | Negative | 9-12mo | Married | No | No | Yes |
| Peter_M | Positive | 18-24mo | Single | No | No | No |
| Caleb_M | Positive | 9-12mo | Married | No | No | Yes |
| Talent_F | Positive | Positive | Single mother | No | No | No |
| Lydia_F | Positive | 18-24mo | Single, never married | No | Yes | Yes |
| Godfrey_M | Positive | 18-24mo | Single, never married | Yes | No | Yes |
| Sipho_M | Positive | 18-24mo | Married | Yes | No | Yes |
| Spiwe_F | Negative | 9-12mo | Married | No | No | No |
| Loveness_F | Negative | 18-24mo | Single mother (1 child) | No | No | Yes |
| Tecla_F | Positive | 18-24mo | Single mother (1 child) | No | No | Yes |
| Nyenge_M | Positive | 9-12mo | Single | Yes | No | Yes |
| Wembe_M | Positive | 9-12mo | Married | No | No | Yes |
| Fanuel_M | Positive | 9-12mo | Married with 2 children | No | No | Yes |

‡=pseudonym and sex of participant

## References

1. World Health Organisation. Global TB Report 2021 [Internet]. Geneva, Switzerland; 2021. Available from: https://www.who.int/publications/i/item/9789240037021

2. World Health Organisation. Global Tuberculosis Report 2020. Geneva, Switzerland; 2020.

3. World Health Organisation. Global Tuberculosis Report 2019 [Internet]. Geneva, Switzerland; 2019. Available from: WHO/CDS/TB/2019.15

4. World Health Organisation. Global Tuberculosis Report 2018 [Internet]. Geneva, Switzerland; 2018. Available from: WHO/CDS/TB/2018.20

5. STOP TB Partnership. The paradigm shift 2016-2020: Global plan to End TB [Internet]. Geneva, Switzerland; 2015. Available from: http://www.stoptb.org/global/plan/plan2/

6. Abidi S, Achar J, Assao Neino MM, Bang D, Benedetti A, Brode S, et al. Standardised shorter regimens versus individualised longer regimens for rifampin-or multidrug-resistant tuberculosis. Eur Respir J [Internet]. 2020;55(1901467):1–13. Available from: http://dx.doi.org/10.1183/13993003.01467-2019

7. University Research Co. LLC. Providing Comprehensive Patient-Centred Care: A conceptual framework for social support for TB patients [Internet]. Bethesda, USA; 2014. Available from: http://www.urc-chs.com/sites/default/files/ProvidingComprehensiveCare.pdf

8. Ghazy RM, Saeh HM El, Abdulaziz S, Hammouda EA, Elzorkany AM, Khidr H. OPEN A systematic review and meta □ analysis of the catastrophic costs incurred by tuberculosis patients. Sci Rep [Internet]. 2022;12(558):1–16. Available from: https://doi.org/10.1038/s41598-021-04345-x

9. Ministry of Health Kenya. The First Kenya Tuberculosis Patient Cost Survey, 2017. Nairobi, Kenya; 2017.

10. Fuady A, Houweling TAJ, Mansyur M, Richardus JH. Catastrophic total costs in tuberculosis-affected households and their determinants since Indonesia ‘ s implementation of universal health coverage. Infect Dis Poverty. 2018;7(3):1–14.

11. Timire C, Ngwenya M, Chirenda J, Metcalfe JZ, Kranzer K, Pedrazzoli D, et al. Catastrophic costs among tuberculosis affected households in Zimbabwe: a national health facility-based survey. Trop Med Int Heal [Internet]. 2021;26(10):1248–55. Available from: https://doi.org/10.1111/tmi.13647

12. Pedrazzoli D, Siroka A, Boccia D, Bonsu F, Nartey K, Houben R, et al. How affordable is TB care? Findings from a nationwide TB patient cost survey in Ghana. Trop Med Int Heal. 2018;23(8):870–8.

13. Barter DM, Agboola SO, Murray MB, Bärnighausen T. Tuberculosis and poverty: the contribution of patient costs in sub-Saharan Africa – a systematic review. BMC Public Health. 2012;980:1–21.

14. Foster N, Vassall A, Cleary S, Cunnama L, Churchyard G, Sinanovic E. The economic burden of TB diagnosis and treatment in South Africa. Soc Sci Med. 2015;130:42–50.

15. Tanimura T, Jaramillo E, Weil D, Raviglione M, Lönnroth K. Financial burden for tuberculosis patients in low- and middle-income countries: a systematic review. Eur Respir J. 2014;43(6):1763–75.

16. Ibarraran P, Regalia F, Stampini M. How Conditional Cash Transfers Work: Good practices after 20 years of implementation. In: Ibarraran P, Regalia F, Stampini M, editors. Inter American Development Bank; 2017. p. 1–8.

17. Food and Agricultural Organisation. Social protection and resilience: Supporting livelihoods in protracted crises and in fragile and humanitarian contexts. Geneva, Switzerland; 2017.

18. Lutge E, Wiysonge CS, Knight SE, Sinclair D, Volmink J. Incentives and enablers to improve adherence in tuberculosis (Review). Cochrane Collab. 2015;(9).

19. Boccia D, Pedrazzoli D, Wingfield T, Jaramillo E, Lönnroth K, Lewis J, et al. Towards cash transfer interventions for tuberculosis prevention, care and control□: key operational challenges and research priorities. BMC Infect Dis [Internet]. 2016;16(307):1–12. Available from: http://dx.doi.org/10.1186/s12879-016-1529-8

20. Rudgard WE, Evans CA, Sweeney S, Wingfield T, Lonnroth K, Barreira D, et al. Comparison of two cash transfer strategies to prevent catastrophic costs for poor tuberculosis-affected households in low- and middle-income countries□: An economic modelling study. PLoS Med. 2017;14(11):1–28.

21. United Nations Development Programme. Discussion paper: Cash Transfers and HIV Prevention. New York, USA; 2014.

22. Grede N, Claros JM, de Pee S, Bloem M. Is There a Need to Mitigate the Social and Financial Consequences of Tuberculosis at the Individual and Household Level□? AIDS Behav. 2014;18:542–53.

23. Lutge E, Lewin S, Volmink J, Friedman I, Lombard C. Economic support to improve tuberculosis treatment outcomes in South Africa□: a pragmatic cluster-randomized controlled trial. Trials [Internet]. 2013;14(154):1–13. Available from: 10.1186/1745-6215-14-154

24. Wingfield T, Boccia D, Tovar MA, Huff D, Montoya R, Lewis JJ, et al. Designing and implementing a socioeconomic intervention to enhance TB control□: operational evidence from the CRESIPT project in Peru. BMC Public Health. 2015;15(810):1–16.

25. World Health Organisation. Global TB Report 2020: Country profiles [Internet]. Geneva, Switzerland; 2020. Available from: https://www.who.int/docs/default-source/hq-tuberculosis/global-tuberculosis-report-2020/country-profile-2020-final-web-min.pdf?sfvrsn=b4137a1c_0

26. Ministry of Health and Child Care. National Tuberculosis Control Programme: Programmatic Management of Drug Resistant Tuberculosis Guidelines. Harare, Zimbabwe; 2014.

27. First Capital Bank Zimbabwe Limited. Forex Reports [Internet]. 2020 [cited 2022 Feb 28]. Available from: https://firstcapitalbank.co.zw/resources/forex-reports/

28. Braun V, Clarke V. Using thematic analysis in psychology. 2008;(March 2013):37–41.

29. Klein K, Bernachea MP, Id SI, Gibbons L, Chirico C, Id FR. Evaluation of a social protection policy on tuberculosis treatment outcomes□: A prospective cohort study [Internet]. Vol. 9. 2019. p. 1–16. Available from: http://dx.doi.org/10.1371/journal.pmed.1002788

30. Reis-santos B, Shete P, Bertolde A, Sales CM, Sanchez MN, Arakaki-sanchez D, et al. Tuberculosis in Brazil and cash transfer programs□: A longitudinal database study of the effect of cash transfer on cure rates. PLoS One [Internet]. 2019;14(2):1–18. Available from: http://dx.doi.org/10.1371/journal.pone.0212617

31. Martins N, Morris P, Kelly PM. Food incentives to improve completion of tuberculosis treatment: randomised controlled trial in Dili, Timor-Leste. Br Med J. 2009;339:b4248.1–8.

32. Carter J, Daniel R, Torrens A, Sanchez M, Marciel E, Bartholomay P, et al. The impact of a governmental cash transfer programme on tuberculosis cure rate in Brazil: A quasi-experimental approach. BMJ Glob Heal. 2019;4(e001029.):1–10.

33. Torrens AW, Rasella D, Boccia D, Maciel ELN, Nery JS. Europe PMC Funders Group Effectiveness of a conditional cash transfer programme on TB cure rate□: a retrospective cohort study in Brazil. 2018;110(242):199–206.

34. Lutge E, Lewin S, Volmink J. Economic support to improve tuberculosis treatment outcomes in South Africa□: a qualitative process evaluation of a cluster randomized controlled trial. Trials. 2014;15(236):1–12.

35. Nirgude AS, Kumar AM V, Timire C, Naik R, Parmar M, Tao L, et al. ‘ I am on treatment since 5 months but I have not received any money ‘: coverage, delays and implementation challenges of ‘ Direct Benefit Transfer ‘ for tuberculosis patients – a mixed-methods study from South India. Glob Health Action [Internet]. 2019;12(1):1– 12. Available from: https://doi.org/10.1080/16549716.2019.1633725

36. Ramma L, Cox H, Wilkinson L, Foster N, Cunnama L, Vassall A, et al. Patients’ costs associated with seeking and accessing treatment for drug-resistant tuberculosis in South Africa. Int J Tuberc Lung Dis [Internet]. 2015;19(12):1513–9. Available from: http://dx.doi.org/10.5588/ijtld.15.0341

37. Boccia D, Bond V. The catastrophic cost of tuberculosis: advancing research and solutions. Int J Tuberc Lung Dis [Internet]. 2019;23(11):1129–30. Available from: http://dx.doi.org/10.5588/ijtld.19.0521

38. Yellappa V, Lefèvre P, Battaglioli T, Narayanan D, Van Der Stuyft P. Coping with tuberculosis and directly observed treatment: a qualitative study among patients from South India. BMC Health Serv Res [Internet]. 2016;16:1–11. Available from: http://dx.doi.org/10.1186/s12913-016-1545-9

39. Rudgard WE, Carter DJ, Scuffell J, Cluver LD, Fraser-hurt N, Boccia D. Cash transfers to enhance TB control□: lessons from the HIV response. 2018;1–7.

40. Meghji J, Gregorius S, Madan J, Chitimbe F, Thomson R, Rylance J, et al. The long term effect of pulmonary tuberculosis on income and employment in a low income, urban setting. Thorax. 2021;76:387–395.

